# SARS-Cov-2 Infection versus Vaccine-Induced Immunity among Veterans

**DOI:** 10.1101/2021.09.27.21264194

**Authors:** Yinong Young-Xu, Jeremy Smith, Caroline Korves

**Affiliations:** White River Junction Veterans Affairs Medical Center, White River Junction, VT; Geisel School of Medicine at Dartmouth, Hanover, NH

## Abstract

**Background:** With over 40 million cases of SARS-CoV-2 infection reported in the US and discussion of both vaccine mandates as well as boosters ongoing, we aim to examine protection conferred by previous infection compared with vaccination so that citizens and policy makers can make informed decisions.

**Objectives:** To compare mRNA COVID-19 vaccine-induced immunity against immunity induced by previous infection with SARS-CoV-2 between June and August 2021 when the Delta variant became dominant in the US.

We conducted a retrospective observational study comparing two groups whose incident vaccination or infection occurred within the first two months of 2021: (1) SARS-CoV-2-naive individuals who received a full mRNA vaccination - 2 doses of either Pfizer or Moderna vaccine, (2) newly infected individuals who were subdivided into those have not been vaccinated and those have been vaccinated after their infection. Matched multivariable Cox proportional hazards model was applied. We evaluated laboratory (RT-PCR) confirmed SARS-CoV-2 infection during follow-up, COVID-related hospitalization, and deaths.

**Setting:** Veterans Health Administration (VHA).

**Main outcomes:** Positive SARS-CoV-2 PCR test, COVID-related hospitalization, and deaths. Protection was estimated from hazard ratios with 95% confidence intervals (CI).

**Results:** A total of 9,539 patients with SARS-CoV-2 infection during the first two months of 2021 were matched to 14,458 and 23,105 patients fully vaccinated with Moderna and Pfizer mRNA vaccines, during the same two months. 3,917 (41%) of patients with SARS-CoV-2 infection were subsequently vaccinated. We plan to study this group separately. Consequently, protections were estimated among those with infection but were not subsequently vaccinated and those vaccinated with a mRNA vaccine. Among seniors, Moderna and Pfizer mRNA vaccines offered stronger protection against infection, lowering the risk by an additional 66% [HR: 0.34 (95% CI, 0.14-0.78)] and 68% [HR: 0.32 (95% CI, 0.14-0.70)]; stronger protection against hospitalization, lowering the risk by an additional 61% [HR: 0.34 (95% CI, 0.14-0.78)] and 45% [HR: 0.34 (95% CI, 0.14-0.78)]; and stronger protection against deaths lowering the risk by an additional 95% [HR: 0.05 (95% CI, 0.004-0.62)] and 99% [HR: 0.01 (95% CI, 0.001-0.44)]. Among young adults (age < 65), the protections offered by vaccines were statistically equivalent to that provided by previous infection, especially in terms of absolute incidence rate.

**Conclusions:** Among the elderly (age 65 or older), two-dose mRNA vaccines provided stronger protection against infection, hospitalization, and death, compared to natural immunity. Among young adults (age < 65), the protections offered between natural immunity and vaccine-induced immunity were similar.

## Introduction

On September 9^th^, 2021, President Biden issued an executive order, requiring COVID-19 vaccinations for federal employees^1^. One week later, a Food and Drug Administration advisory panel overwhelmingly voted in favor of giving Pfizer-BioNTech’s Covid-19 booster vaccinations to people 65 years and older as well as those at high risk of severe illness^2^. Prior to these actions, an Israeli research group found that “natural immunity confers longer lasting and stronger protection against infection, symptomatic disease and hospitalization caused by the Delta variant of SARS-CoV-2, compared to the BNT162b2 two-dose vaccine-induced immunity”^3^. Their finding was alarming: never-infected people who were vaccinated in January and February were, six to 13 times more likely to get infected than unvaccinated people who were previously infected with the coronavirus, in June, July, and the first half of August. Because their study population included mostly young adults, with an average age of 36, and because Moderna COVID-19 vaccine was not studied, we aimed to validate their findings in the US Veterans population where half are 65 or older^4^ and both mRNA vaccines were available.

## Methods

The study protocol was approved by the institutional review board of the VA Medical Center in White River Junction, VT and was granted a waiver of consent. The authors followed STROBE reporting guidelines.

### Data Source

The Veterans Health Administration (VHA) is the largest integrated health care system in the U.S., providing comprehensive care to over nine million Veterans at more than 171 medical centers and 1,112 outpatient sites of care^5^. Electronic health record data from the VHA Corporate Data Warehouse (CDW) were analyzed. The CDW contains patient-level information on all patient encounters, treatments, prescriptions (including vaccinations), and laboratory results rendered in VHA medical facilities. CDW data are also linked to other patient-level data, including: 1) demographic and mortality data from the VHA Enrollee Status File, which incorporates data from external sources including the Social Security Administration and Veterans Benefits Administration, and 2) claims data on VHA enrollees from Medicare fee-for-service files.

### Study design and population

We conducted a retrospective cohort study, using comprehensive testing and vaccination data from the CDW. To make our results comparable, we matched part of study design to that of Gazit et al^3^, focusing on June, July, and the first half of August. The study population included all Veterans enrolled under the care of VHA aged 18 or older. Like the study by Gazit et al, patients were SARS-CoV-2-naïve prior to January 1, 2021, and then were either fully vaccinated or had a documented, laboratory-confirmed, SARSCoV-2 infection prior to March 1, 2021. The 2^nd^ dose vaccination date or specimen collection date for infection during these two months was the index date.

Each previously infected Veteran was matched with up to four vaccinated individuals on the following: state and index event dates (within +/-2 weeks), race/ethnicity, age groups, sex, rural/urban, CCI, and VHA priority. A three-month minimum interval was required between the index date and the events of interest (positive SARS-CoV-2 test identified by RT-PCR, hospitalization, and death), following the 90-day guideline of the CDC to distinguish between viral shedding of one infection and a new episode. This also incidentally helped to focus on COVID-19 infections during June through August, a period where the predominant variant in the US was Delta. Follow-up time started at the index date, included a minimum 90-day interval where no events were measured as outcomes, and ended on the date of the earliest positive test of SARS-CoV-2 infection, death, or the end of the study period (August 18^th^, 2021).

### Baseline Characteristics

Characteristics measured during the baseline period (i.e., 1 year prior to study entry) included demographics and comorbidities. Demographics included age, sex, race, rurality, and priority rating for VHA care as a proxy for socioeconomic status^6^. We examined the presence of clinical comorbidities within 12 months preceding hospitalization using ICD-10 diagnosis codes recorded in patients’ electronic medical records and categorized them according to the Charlson Comorbidity Index (CCI). The CCI score is a validated, weighted measure that predicts one-year mortality, with higher scores indicating greater illness burden^7^. Additionally, we reported the top comorbidities including cardiovascular diseases, hypertension, diabetes, chronic kidney disease, chronic obstructive pulmonary disease, immunocompromised conditions, and cancer.

### Statistical Analysis

We constructed matched Cox survival models to compare time to events of interest by type of immunity.

All tests were two-tailed, and 0.05 was the chosen level of statistical significance. Data and baseline characteristics were constructed using SAS 9.4 (SAS Institute, Cary, North Carolina). Modeling was performed using Stata 17 (StataCorp, College Station, Texas).

## Results

A total of 9,539 patients with SARS-CoV-2 infection during the first two months of 2021 were matched to 14,458 and 23,105 participants fully vaccinated with Moderna and Pfizer mRNA vaccines, respectively. (Table 1) Between June and August, a total of 110 (0.23%) participants tested positive for COVID-19 (Table 2) with those previously infected without subsequent vaccination having the highest infection rate – 2.7 per 100,000 patient-days. These data we further analyzed by age with 65 as the cutoff. Among those 65 or older, those previously infected had the highest infection rate, 4.8 per 100,000 patient-days, followed by Pfizer at 1.5, and Moderna, 1.2. Matched, adjusted multivariable Cox model showed that, with previously infected patients as reference, those received Moderna and Pfizer vaccines had 66% [HR: 0.34 (95% CI, 0.14-0.78)] and 68% [HR: 0.32 (95% CI, 0.14-0.70)] significantly lower risk of infection between June and August. (Table 3)

**Table 1.**
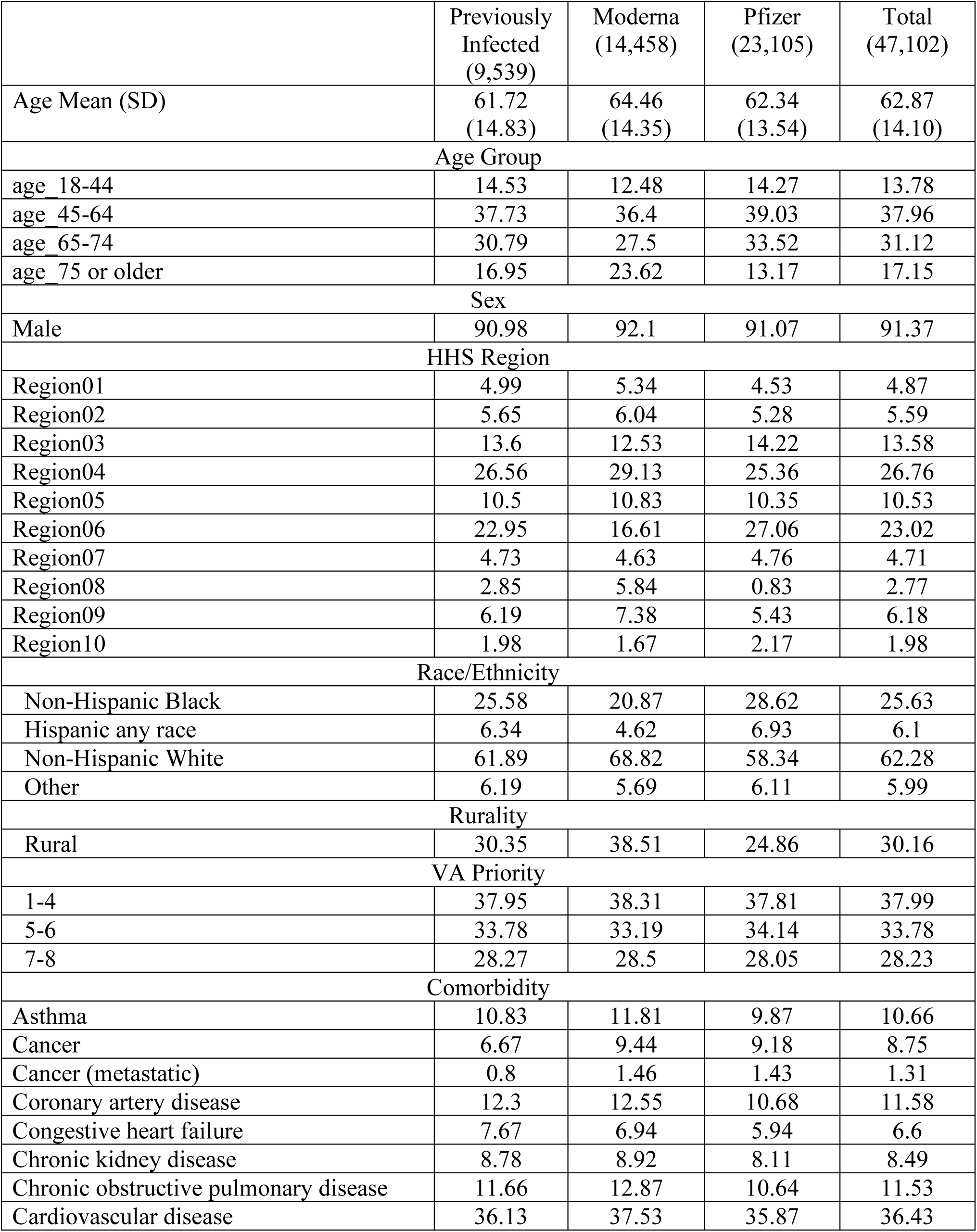
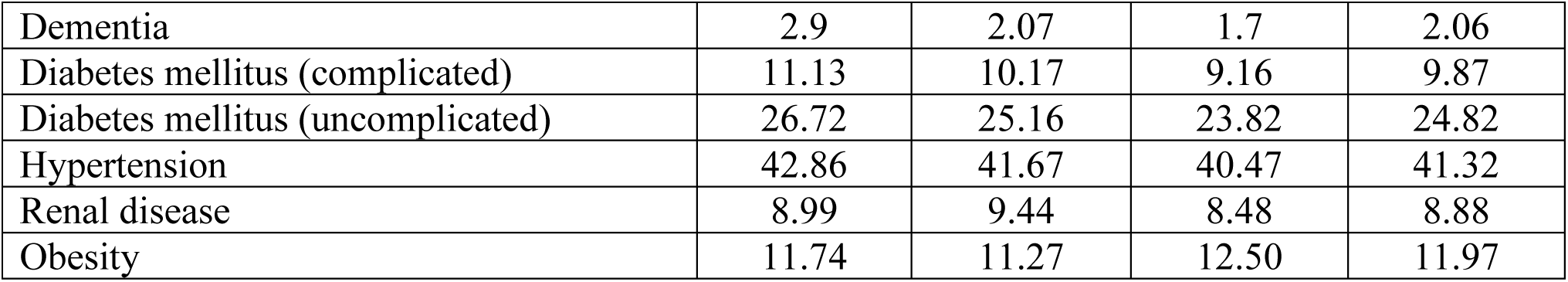
Exposure Status and Baseline Characteristics of Study Subjects (%)

**Table 2.** Index events between 1/1/21 and 3/1/21, Number of Covid-19 infection

| Index Event | Subjects | June | July | August | Total | proportion | Incidence Rate (per 1000-person 100 day) |
| --- | --- | --- | --- | --- | --- | --- | --- |
| Infections (positive PCR test) |  |  |  |  |  |  |  |
| Moderna | 14,458 | 2 | 8 | 15 | 25 | 0.17% | 0.9 |
| Pfizer | 23,105 | 3 | 25 | 29 | 57 | 0.25% | 1.3 |
| Infected | 9,539 | 20 | 5 | 3 | 28 |  |  |
| - vaccine | 5,622 | 20 | 5 | 3 | 28 | 0.50% | 2.7 |
| + vaccine | 3,917 | - | - | - | - | - | - |
| Age 65+ |  |  |  |  |  |  |  |
| Moderna | 7,391 | 2 | 6 | 8 | 16 | 0.22% | 1.2 |
| Pfizer | 10,789 | 2 | 11 | 17 | 30 | 0.28% | 1.5 |
| Infected | 2,480 | 15 | 2 | 2 | 19 | 0.77% | 4.8 |
| Age < 65 |  |  |  |  |  |  |  |
| Moderna | 7,067 | 0 | 2 | 7 | 9 | 0.13% | 0.7 |
| Pfizer | 12,316 | 1 | 14 | 12 | 27 | 0.22% | 1.2 |
| Infected | 3,142 | 5 | 3 | 1 | 9 | 0.29% | 1.4 |
| Hospitalization |  |  |  |  |  |  |  |
| Age 65+ |  |  |  |  |  |  |  |
| Moderna | 7,391 | 10 | 8 | 12 | 30 | 0.41% | 2.2 |
| Pfizer | 10,789 | 13 | 18 | 18 | 49 | 0.45% | 2.5 |
| Infected | 2,480 | 7 | 5 | 1 | 13 | 0.52% | 3.2 |
| Age < 65 |  |  |  |  |  |  |  |
| Moderna | 7,067 | 10 | 9 | 11 | 30 | 0.42% | 2.3 |
| Pfizer | 12,316 | 22 | 26 | 11 | 59 | 0.48% | 2.9 |
| Infected | 3,142 | 2 | 4 | 0 | 6 | 0.19% | 0.9 |
| Death |  |  |  |  |  |  |  |
| Age 65+ |  |  |  |  |  |  |  |
| Moderna | 7,391 | 0 | 1 | 0 | 1 | 0.01% | 0.1 |
| Pfizer | 10,789 | 0 | 0 | 4 | 4 | 0.04% | 0.2 |
| Infected | 2,480 | 2 | 0 | 0 | 2 | 0.08% | 0.5 |
| Age < 65 |  |  |  |  |  |  |  |
| Moderna | 7,067 | 1 | 0 | 0 | 1 | 0.01% | 0.08 |
| Pfizer | 12,316 | 0 | 1 | 1 | 2 | 0.02% | 0.09 |
| Infected | 3,142 | 0 | 0 | 0 | 0 | 0 | 0 |

**Table 3.** Risk [HR (95% CI)] of COVID-19 infection (PCR test positive), by age and mRNA vaccines (Reference Group are those previously infected)

| Variable Periods |  |  | June-August |  |
| --- | --- | --- | --- | --- |
| Age Group | <65 | 65+ | <65 | 65+ |
| Infection (July, August) |  |  | Infection |  |
| Moderna | 1.04 (0.24-4.58) | 2.45 (0.56-20.66) | 0.35 (0.11, 1.13) | 0.34 (0.14,0.78) |
| Pfizer | 1.59 (0.41-6.11) | 1.89 (0.49-7.28) | 0.64 (0.24, 1.69) | 0.32 (0.14,0.70) |
| Hospitalization (April-August) |  |  | Hospitalization |  |
| Moderna | 0.85 (0.43-1.71) | 0.39 (0.20-0.80) | 1.46 (0.55-3.88) | 0.76 (0.31-1.83) |
| Pfizer | 1.21 (0.64-2.28) | 0.55 (0.29-1.05) | 2.33 (0.92-5.93) | 0.82 (0.36-1.88) |
| Death (April-August) |  |  | Death |  |
| Moderna | N/A | 0.05 (0.004-0.62) | N/A | 0.70 (0.04-11.79) |
| Pfizer | N/A | 0.01 (0.001-0.44) | N/A | N/A |
| Hospitalization and Mortality (April-August) |  |  | Hospitalization and Mortality |  |
| Moderna | 0.88 (0.43-1.72) | 0.31 (0.16-0.59) | 1.47 (0.55-3.90) | 0.75 (0.32-1.73) |
| Pfizer | 1.20 (0.64-2.26) | 0.40 (0.22-0.71) | 2.30 (0.91-5.83) | 0.70 (0.32-1.53) |
| All combined |  |  | All combined |  |
| Moderna | 0.52 (0.30-0.88) | 0.26 (0.16-0.42) | 0.86 (0.43-1.74) | 0.62 (0.33-1.16) |
| Pfizer | 0.71 (0.44-1.14) | 0.29 (0.19-0.46) | 1.26 (0.66-2.41) | 0.57 (0.32-1.01) |

For those younger than 65, this pattern remained the same. Those previously infected had the highest infection rate, 1.4 per 100,000 patient-days, and those vaccinated with Modern vaccine had the lowest infection rate at 0.7. In between them, those vaccinated with Pfizer vaccine had an infection rate of 1.2%. For these young adults, matched, adjusted multivariable Cox model showed that, again with previously infected patients as reference, those who received Pfizer and Moderna vaccines had 65% [HR: 0.71 (95% CI, 0.11-1.13)] and 36% [HR: 0.64 (95% CI, 0.24-1.69)] lower risk of infection, but no longer statistically significant. If we focus on relative risk and focus on July and August, two months during which the prevalence of Delta variant reached 100% in most of the US, vaccination showed reduced protection in comparison to previous infection against breakthrough infection, although not statistically significant and far smaller than reported by Gazit et al.

Severe outcomes such as hospitalization and death have important societal implications and larger impact on the nation’s healthcare infrastructure. These events are relatively evenly distributed between June and August (Table 2) and extend back to April and May where some of these events occurred at least 90 days after the index event. Here we found that seniors who were vaccinated had 69% [Moderna HR: 0.31 (95% CI, 0.16-0.59)] and 60% [Pfizer HR: 0.40 (95% CI, 0.22-0.71)] significantly lower risk of infection between April and August. (Table 3) Against the composite outcome of hospitalization and death, the pattern of protection was not clear. We found that young adults who were vaccinated had 12% lower [Moderna HR: 0.88 (95% CI, 0.43-1.72)] and 20% higher [Pfizer HR: 1.20 (95% CI, 0.64-2.26)] risk of infection between April and August, but neither was statistically significant. (Table 3)

## Discussion

Contrary to the findings by Gazit et al., our study showed that, among the elderly, two-dose mRNA vaccines provided stronger protection against infection, hospitalization, and death, compared to natural immunity, and not the other way around. Although during June to August, especially during July and August, this impact lessened, but overall, the data showed more protection among mRNA vaccines recipients: [Moderna HR: 0.26 (95% CI, 0.16-0.42)] and [Pfizer HR: 0.29 (95% CI, 0.19-0.46)]. These differences might have resulted from our different approaches such as statistical model (survival vs. logistic regression), population (average age 62 vs. 36), and the benefit of having another mRNA vaccine from Moderna. Moreover, in June, Israel has already reached herd immunity which tends to make those vaccinated and unvaccinated appear similar thus bias towards an underestimation of the effect of vaccination.

We found the relative protection offered by natural immunity vs vaccine-induced immunity differed by age. Although the data showed mostly additional protection by mRNA vaccines among the elderly, among young adults we mostly see less protection by mRNA vaccines. Nevertheless, in terms of absolute infection rates, the protection was similar as those previously infected. (Figure 1) Immunosenescence among older people could impact the ability to respond to infections and maintain long-term immune memory that was acquired either by infection or vaccination. By focusing on July and August, during which the prevalence of Delta variant reached 100% in most of the US, we intended to study the impact of Delta breakthrough infections. Unfortunately, this approach is ecological by nature owing to a paucity of whole genome sequencing data in the US. Individually, vaccine effectiveness has been reported to wane over time and could enter a particularly less effective period after 5 months.

**Figure 1.**
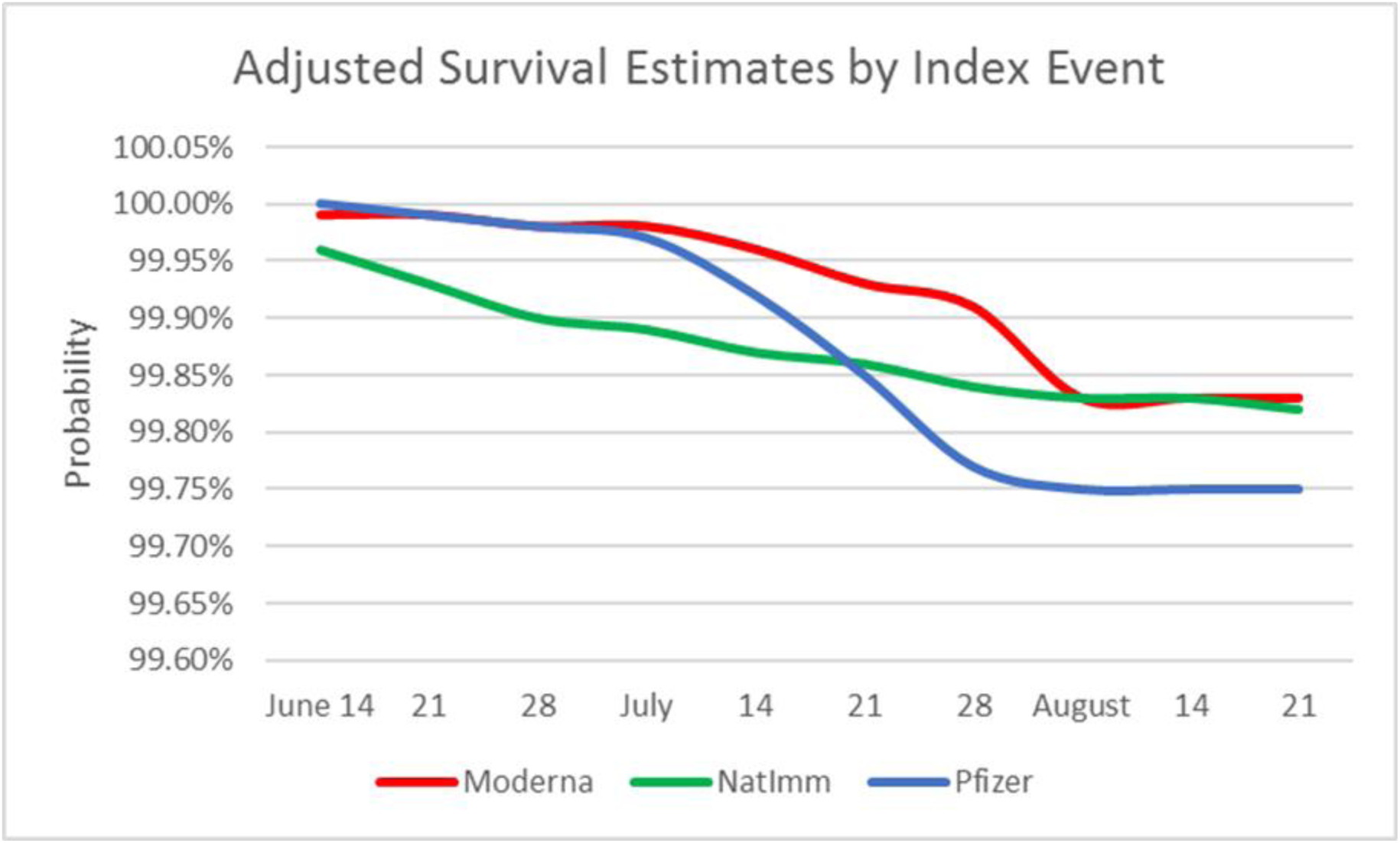

## Limitation

Death is an important competing risk as one must survive the first infection to become at risk again for another infection. The sharp rise in infections among the vaccinated during July and August could be attributed to the impact of Delta, the waning effect of the vaccines, and the relaxation of public health measures in the US starting in May. Most re-infections occurred during the 4^th^ month after previous infection. It is possible that re-infected patients had residual positive PCR rather than new infections. That could change the picture somewhat. (Figure 2)

**Figure 2.**
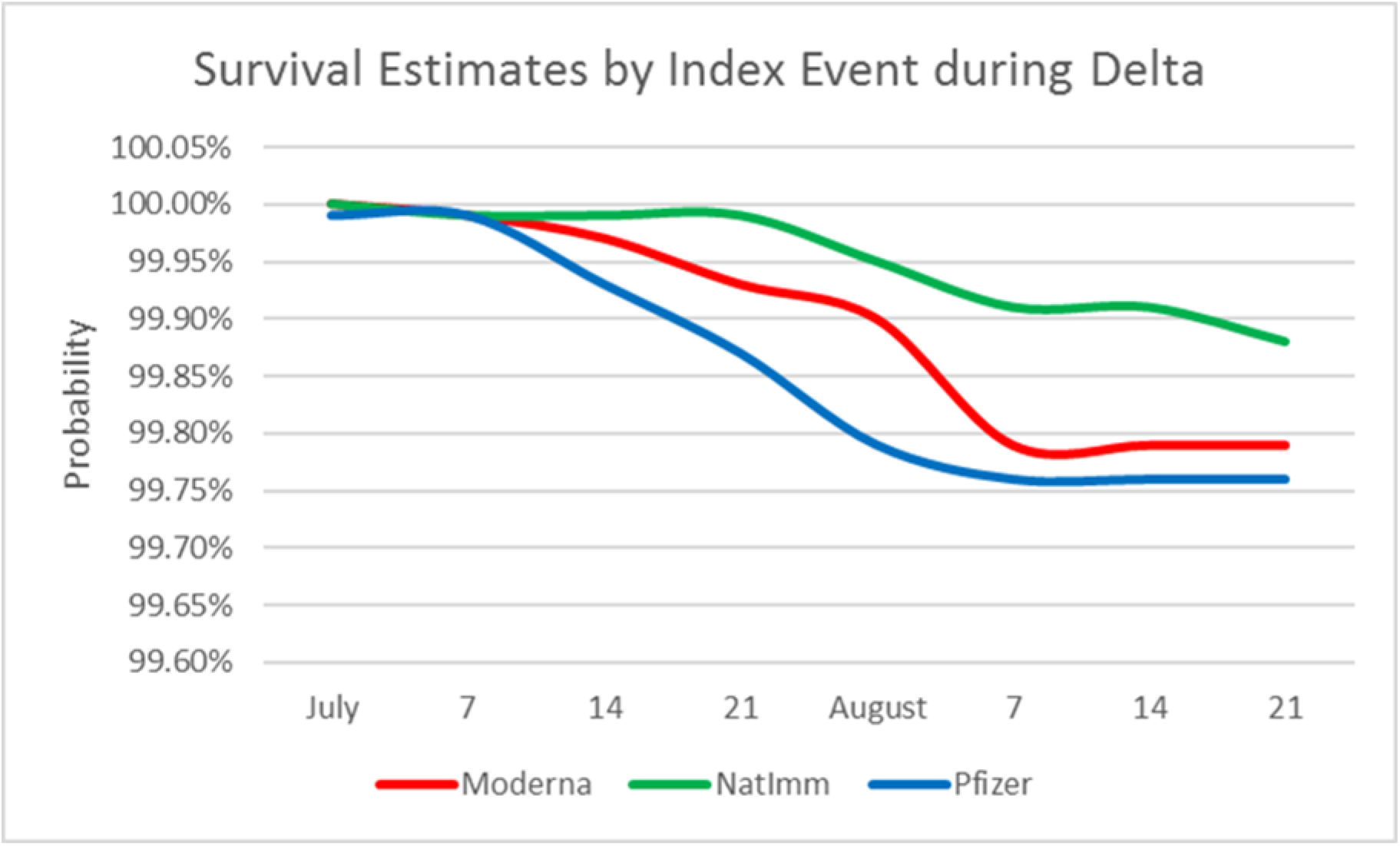

Every study population is unique. Half of our study participants were aged 65 years and older, over 90% were male, and a high prevalence of underlying conditions. During the first two months of 2021, VHA deployed more Pfizer vaccines in urban areas due to refrigeration requirement.

## Conclusion

As of September 2021, there have been 42 million reported cases according to the CDC, and four out of seven (24 million) are aged between 18 and 65. Many of them will face the decision or mandates regarding vaccination. If natural immunity is replied upon for protection against COVID-19 infection among younger adults, then we must study it carefully and follow it continuously to determine its protection relative to natural immunity plus vaccination, and vaccination alone.

## Data Availability

Data are available upon request to the lead author.

## Acknowledgements of research support for the study

This project was funded by the U.S. Department of Veterans Affairs (VA) Office of Rural Health.

Yinong Young-Xu had full access to all the data in the study and takes responsibility for the integrity of the data and the accuracy of the data analysis.

## Reference

1 https://www.whitehouse.gov/briefing-room/presidential-actions/2021/09/09/executive-order-on-requiring-coronavirus-disease-2019-vaccination-for-federal-employees/

2 https://www.nbcnews.com/health/health-news/fda-advisory-group-rejects-covid-boosters-limits-high-risk-groups-rcna2074

3 https://www.medrxiv.org/content/10.1101/2021.08.24.21262415v1.full-text

4 Profile of Veterans: 2014 – Data from the American Community Survey. In: U.S. Department of Veterans Affairs [online]. Available at: www.va.gov/vetdata/docs/SpecialReports/Profile_of_Veterans_2014.pdf. Accessed September 15, 2020.

5 https://www.va.gov/health/

6 Petersen LA, Byrne MM, Daw CN, Hasche J, Reis B, Pietz K. Relationship between clinical conditions and use of Veterans Affairs health care among Medicare-enrolled veterans. Health Serv Res. 2010; 45:762–791. Available at: http://www.doi.org/10.1111/j.1475-6773.2010.01107.x.

7 De Groot V, Beckerman H, Lankhorst GJ, Bouter LM. How to measure comorbidity: a critical review of available methods. J Clin Epidemiol. 2003;56(3):221–229. pmid:12725876

